# Novel mammogram-based measures improve breast cancer risk prediction beyond an established mammographic density measure

**DOI:** 10.1101/2020.05.24.20111815

**Authors:** Tuong L. Nguyen, Daniel F. Schmidt, Enes Makalic, Gertraud Maskarinec, Shuai Li, Gillian S. Dite, Ye K. Aung, Christopher F. Evans, Ho N. Trinh, Laura Baglietto, Jennifer Stone, Yun-Mi Song, Joohon Sung, Robert J. MacInnis, Pierre-Antoine Dugué, James G. Dowty, Mark A. Jenkins, Roger L. Milne, Melissa C. Southey, Graham G. Giles, John L. Hopper

**Affiliations:** Centre for Epidemiology and Biostatistics, Melbourne School of Population and Global Health, University of Melbourne, 207 Bouverie Street, Parkville, Victoria 3010, Australia; Faculty of Information Technology, Monash University, Clayton, Victoria, Australia; University of Hawaii Cancer Center, Honolulu, Hawaii, USA; Genetic Technologies Ltd, 60–66 Hanover St, Fitzroy VIC 3065, Australia; Department of Clinical and Experimental Medicine, University of Pisa, Italy; School of Population and Global Health, University of Western Australia, Perth, Western WA, 6009, Australia; Department of Family Medicine, Samsung Medical Center, Sungkyunkwan University School of Medicine, Irwon-ro 81, Gangnamgu, Seoul, South Korea; Department of Epidemiology School of Public Health, Seoul National University, 1 Gwanak-ro, Gwanak-gu, 151-742 Seoul, South Korea; Institute of Health and Environment, Seoul National University, 1 Gwanak-ro, Gwanak-gu, 151-742 Seoul, South Korea; Cancer Epidemiology Division, Cancer Council Victoria, Melbourne, Victoria, Australia; Precision Medicine, School of Clinical Sciences at Monash Health, Monash University, Clayton, Victoria, Australia

**Keywords:** breast cancer, Cirrus, Cirrocumulus, interval breast cancer, mammographic density, screen-detected breast cancer

## Abstract

Mammograms contain information that predicts breast cancer risk. We developed two novel mammogram-based breast cancer risk measures based on image brightness (*Cirrocumulus*) and texture (*Cirrus*). Their risk prediction when fitted together, and with an established measure of conventional mammographic density (*Cumulus*), is not known. We used three studies consisting of: 168 interval cases and 498 matched controls; 422 screen-detected cases and 1,197 matched controls; and 354 younger-diagnosis cases and 944 controls frequency-matched for age at mammogram. We conducted conditional and unconditional logistic regression analyses of individually- and frequency-matched studies, respectively. We estimated measure-specific risk gradients as the change in odds per standard deviation of controls after adjusting for age and body mass index (OPERA) and calculated the area under the receiver operating characteristic curve (AUC). For interval, screen-detected and younger-diagnosis cancer risks, the best fitting models (measure-specific OPERAs [95% confidence intervals]) involved: *Cumulus* (1.81 [1.41 to 2.31]) and *Cirrus* (1.72 [1.38 to 2.14]); *Cirrus* (1.49 [1.32 to 1.67]) and *Cirrocumulus* (1.16 [1.03 to 1.31]); and *Cirrus* (1.70 [1.48 to 1.94]) and *Cirrocumulus* (1.46 [1.27 to 1.68]), respectively. The AUCs were: 0.73 [0.68 to 0.77], 0.63 [0.60 to 0.66], and 0.72 [0.69 to 0.75], respectively. Combined, our new mammogram-based measures doubled the risk gradient for screen-detected and younger-diagnosis breast cancer (P<10^−12^), have at least the same discriminatory power as the current polygenic risk score, and are more correlated with causal factors than conventional mammographic density. Discovering more information about breast cancer risk from mammograms could help enable risk-based personalised breast screening.

**What’s new?:** We developed two novel mammogram-based breast cancer risk measures based on image brightness (Cirrocumulus) and texture (Cirrus). We estimated their risk prediction when fitted with conventional mammographic density (Cumulus), for interval, screen-detected, and younger age at diagnosis breast cancer. Our new measures substantially improved risk prediction. There is more risk information in a woman’s mammogram than in her genome. Discovering new ways of extracting risk information from a mammogram could enable risk-based personalised breast screening.

## INTRODUCTION

Historically, mammographic density has been defined as the light *or* bright areas on a mammogram (we call this *Cumulus*) and has well-established risk associations with breast cancer overall and with both interval and screen-detected cancers [1]. But there has been debate about the extent to which these associations are due to existing tumours being missed at mammographic screening, especially for interval cancers. It is also not clear if mammographic density, defined as above or otherwise, or other mammogram-based measures, are the best predictors of breast cancer.

We addressed these issues by trying to discover aspects of a mammographic image that differ between women with and without breast cancer. First, we redefined mammographic density at, in effect, a higher pixel brightness threshold to encompass just the brightest regions to create *Cirrocumulus* [2-5]. Second, we applied machine learning to textural patterns to create *Cirrus* [6].

We had previously considered each new measure separately with the established measure. Both *Cirrus* and *Cirrocumulus* were correlated with *Cumulus*; r∼0.4 and 0.6, respectively [2-6]. Except for interval cancer, we found that, when fitted together, the *Cumulus* risk gradient substantially decreased compared with being fitted alone. On the other hand, the *Cirrocumulus* and *Cirrus* risk gradients both remained similar to what they were when fitted alone [2-6]. We concluded that “conventional mammographic density predicts interval cancer due to its role in masking, while the new mammogram-based risk measures could have a causal effect on both interval and screen-detected breast cancer” [7].

We had assessed the strength of risk prediction, in terms of the ability to differentiate cases from controls on a population basis, using the change in odds per adjusted standard deviation (OPERA) [8]. OPERA uses the standard deviation of the residuals for controls after adjusting the mean for factors such as age and body mass index, not the standard deviation of the cross-sectional distribution of the raw measure itself. The resulting risk gradients, therefore, take into account the negative confounding of age and body mass index.

In terms of interpretation, the residuals are in effect the risk factor (not the raw measure itself), and the log(OPERA) is the difference between cases and controls in the mean of the standardised residuals; see Appendix [6]. On the other hand, the OR per *unadjusted* standard deviation is problematic when it is estimated after *adjusting* for covariates (unless it is adjusted in a way that depends on the correlations between covariates [2,7]) and is prone to misinterpretation; see Discussion.

In this paper, we aimed to determine the extent to which our new measures are correlated with one another, and the extents to which risk prediction is improved when our new measures are fitted together with and without also fitting an established mammographic density measure.

We have again used the OPERA approach because it allows multiple correlated risk factors to be compared and put into perspective in terms of risk discrimination in ways that are not possible using, for example, *change* in the area under the receiver operating characteristic curve (AUC), which depends on the order in which the factors are included. Individually, our new mammogram-based risk measures are among the strongest of all currently known breast cancer risk factors [6]. But it is not known what risk prediction is obtained when these are fitted together with the conventional mammographic density measure.

We also used our findings to see how the risk predictions obtained from combining our new measures compare with that which would be achieved by other breast cancer risk factors, including polygenic risk scores.

## MATERIAL AND METHODS

### Sample

We used data from three independent studies: (i) a nested case-control study of 168 cases with interval breast cancer (those diagnosed within two years of a negative screen) and 498 matched controls within the Melbourne Collaborative Cohort Study (MCCS) [5, 9-11]; (ii) a nested case-control study of 422 cases with screen-detected breast cancer and 1,197 matched controls within the MCCS [5, 9-10]; and (iii) a case-control study of 354 cases with on average younger-diagnosis breast cancers and 944 controls from the Australian Breast Cancer Family Study and the Australian Mammographic Density Twins and Sisters Study, both over-sampled for early-onset disease or having a family history of breast cancer, and frequency matched for age at mammogram in 5-year age groups, and for family history [3]. For study (iii), the age at mammogram was under 45 years for 40% of cases and 39% of controls, respectively, and by design, 30% of cases and 29% of controls had a family history of breast cancer compared with 10% of controls in studies (i) and (ii).

Table 1 show that, for all studies, the average ages at diagnosis of the cases and the average ages at mammogram of the controls were similar. The mean (standard deviation) age at diagnosis was 62.3 (7.3) years for the interval cancers, 64.3 (8.2) years for the screen-detected cancers, and 48.5 (10.7) years for the younger-diagnosis cases. For the prospective studies (i) and (ii), the average times between mammogram and diagnosis were 5 and 6 years (standard deviation 3) for interval and screen-detected cancers, respectively. For study (iii), for 68% of the affected women we used the mammogram at diagnosis from the side opposite to that in which the cancer was diagnosed, otherwise we used mammograms taken closest before diagnosis (by on average 4 years). For the unaffected women, we studied the mammogram of a randomly chosen breast. All participants gave written informed consent and the studies were approved by appropriate human research ethics committees.

**Table 1.**
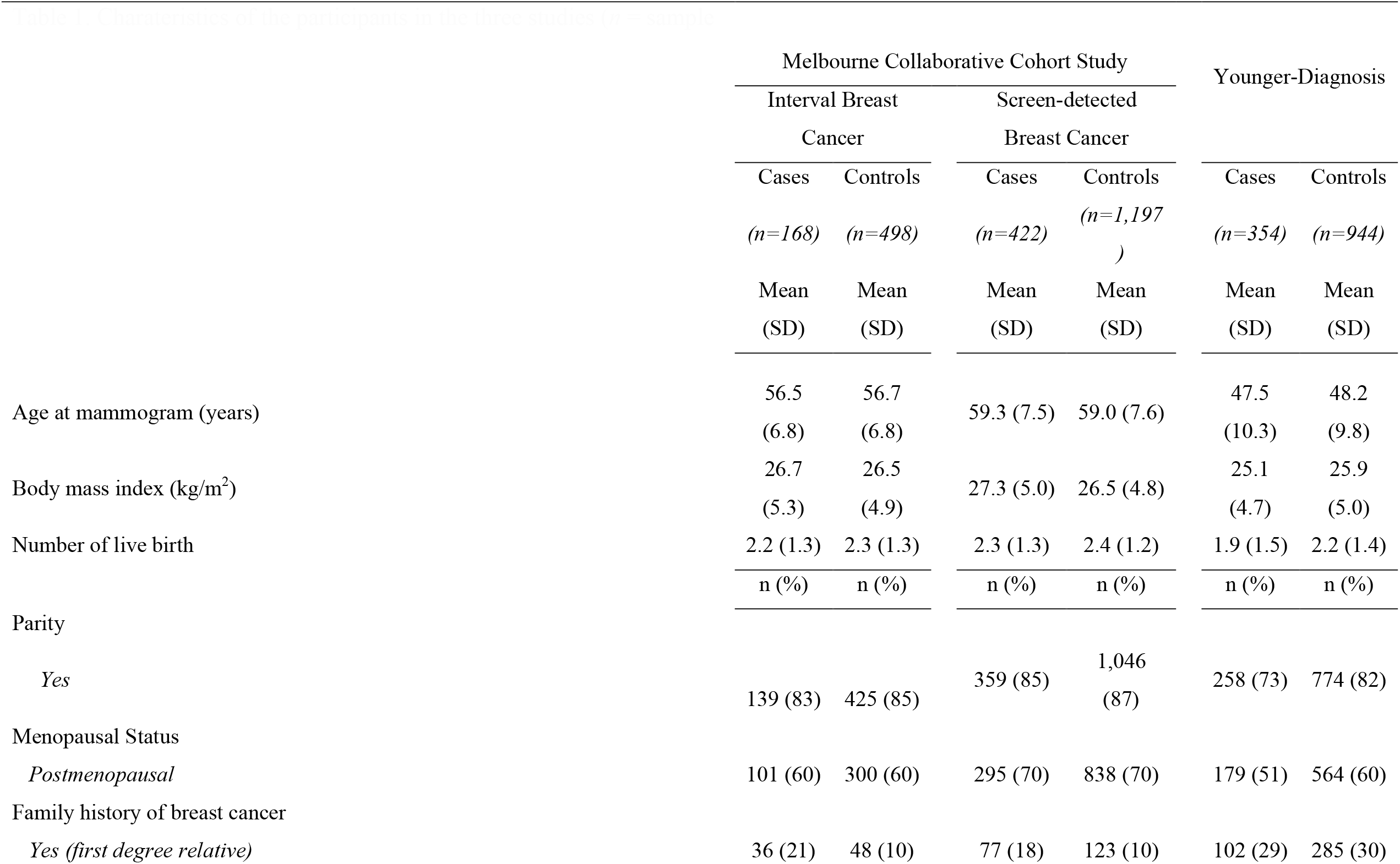
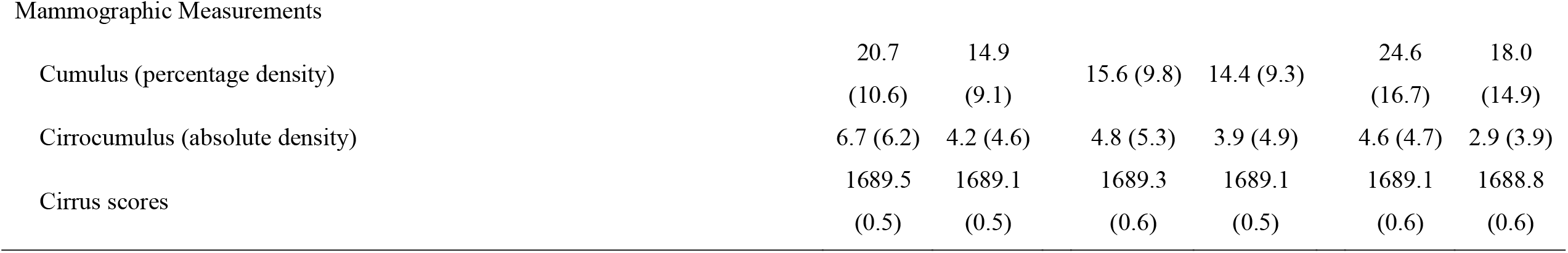
Charateristics of the participants in the three studies (*n* = sample size; SD = standard deviation)

|  | Melbourne Collaborative Cohort Study |  |  |  | Younger-Diagnosis |  |
| --- | --- | --- | --- | --- | --- | --- |
|  | Interval Breast |  | Screen-detected |  |  |  |
|  | Cancer |  | Breast Cancer |  |  |  |
|  | Cases | Controls | Cases | Controls | Cases | Controls |
|  | (*n*=168) | (*n*=498) | (*n*=422) | (*n*=1,197) | (*n*=354) | (*n*=944) |
|  | Mean | Mean | Mean | Mean | Mean | Mean |
|  | (SD) | (SD) | (SD) | (SD) | (SD) | (SD) |
| Age at mammogram (years) | 56.5<br>(6.8) | 56.7<br>(6.8) | 59.3 (7.5) | 59.0 (7.6) | 47.5<br>(10.3) | 48.2<br>(9.8) |
| Body mass index (kg/m<sup>2</sup>) | 26.7<br>(5.3) | 26.5<br>(4.9) | 27.3 (5.0) | 26.5 (4.8) | 25.1<br>(4.7) | 25.9<br>(5.0) |
| Number of live birth | 2.2 (1.3) | 2.3 (1.3) | 2.3 (1.3) | 2.4 (1.2) | 1.9 (1.5) | 2.2 (1.4) |
|  | n (%) | n (%) | n (%) | n (%) | n (%) | n (%) |
| Parity |  |  |  |  |  |  |
| *Yes* | 139 (83) | 425 (85) | 359 (85) | 1,046<br>(87) | 258 (73) | 774 (82) |
| Menopausal Status |  |  |  |  |  |  |
| *Postmenopausal* | 101 (60) | 300 (60) | 295 (70) | 838 (70) | 179 (51) | 564 (60) |
| Family history of breast cancer |  |  |  |  |  |  |
| *Yes (first degree relative)* | 36 (21) | 48 (10) | 77 (18) | 123 (10) | 102 (29) | 285 (30) |
| Mammographic Measurements |  |  |  |  |  |  |
| Cumulus (percentage density) | 20.7<br>(10.6) | 14.9<br>(9.1) | 15.6 (9.8) | 14.4 (9.3) | 24.6<br>(16.7) | 18.0<br>(14.9) |
| Cirrocumulus (absolute density) | 6.7 (6.2) | 4.2 (4.6) | 4.8 (5.3) | 3.9 (4.9) | 4.6 (4.7) | 2.9 (3.9) |
| Cirrus scores | 1689.5<br>(0.5) | 1689.1<br>(0.5) | 1689.3<br>(0.6) | 1689.1<br>(0.5) | 1689.1<br>(0.6) | 1688.8<br>(0.6) |

### Mammogram-based measures

Using digitised film mammograms, the *Cumulus* and *Cirrocumulus* measures were created using the computer-assisted threshold software CUMULUS [12] and the *Cirrus* measures were created as previously described [6]. Each density measure was first transformed using the Box–Cox power transformation to have an approximately normal distribution. The appropriate transformations were cube-root for the *Cumulus* and logarithm for *Cirrocumulus. Cirrus* was left untransformed.

Using the controls, the means of the transformed measures were adjusted for age and body mass index as linear terms, and scaled by the standard deviation of the residuals. This parameterisation of the mean, derived using the controls, was used to derive the residuals both for the controls and for the cases. These residuals were then standardised by dividing by the standard deviation of the residual for the controls. These standardised residuals, called *Cumulus, Cirrocumulus* and *Cirrus*, were then fitted in the analyses.

For *Cumulus* we used the percentage measure because it substantially better predicted interval breast cancer (difference in log likelihood 14; see also [10]). and because for screen-detected and younger-diagnosis cancer the results did not differ when we used absolute *Cumulus* (data not shown). For *Cirrocumulus* we used the absolute measure because it has less measurement error by not having to be divided by total area and then adjusted for BMI, which is correlated with total area.

### Statistical Methods

All analyses were conducted using the Stata software [13]. For descriptive purposes, we presented the numbers of cases and controls for each pair of measures, and the estimated risks relative to the average risk for the population from which the controls were sampled for the different tertile-by-tertile categories (based on controls) using the cci option, which compares the case-control frequencies in each category with the case-control frequency overall.

The control data was used to investigate the joint distributions of the pairs of measures and to estimate the proportion of the relevant population in the different categories.

Risk gradients in terms of OPERAs for the measures *Cumulus, Cirrocumulus* and *Cirrus* (see Mammogram-based measures) were estimated using conditional logistic regression for the two nested matched case-control studies and using unconditional logistic regression for the frequency-matched case-control study. To compare fits and select the best fitting models, we used the likelihood ratio criterion [14] with *P*<0.05 considered to be the threshold for nominal statistical significance. We also used the likelihood ratio test to assign statistical significances to the changes in AUC between nested models. We estimated log(OPERA) as a function of age and BMI, and family history and used the likelihood ratio criterion to test for differences. For each measure, we used the estimates of log(OPERA) and their standard errors to conduct statistical tests of differences across the three studies under the assumption of independence.

The risk gradient, and hence the ability to differentiate cases from controls on a population basis, was reported as the OPERA for which we used adjustment for age at mammogram and body mass index [8]. We present OPERA estimates in the tables for ease of interpretation and log(OPERA) in the text because it is a natural scale on which to assess risk gradients. Under the assumptions of a multiplicative risk model for a normally distributed risk factor, log(OPERA) = √2Φ^−1^(AUC), where Φ is the normal (0,1) distribution function (see Supplementary Material in [6]), so that the AUC is approximately linearly related to log(OPERA), at least in the range of AUC from 0.5 to 0.7 (OPERA from 1 to 2). Under the multiplicative risk model, log(OPERA) is equal to the difference in means between cases and controls divided by the standard deviation of the adjusted risk factor; the inter-quartile risk ratio is approximately OPERA^2.5^. When we fitted multiple risk measures together, we used the AUC and the formula above to calculate the AUC-equivalent OPERA as if we had fitted one combined measure.

As a comparison of analytic approaches, we fitted the transformed but unadjusted measures (rather than the standardised residuals after adjustment for age and BMI). To distinguish this, we call them Percent Density, Cirrocumulus and Cirrus and presented the odds ratio per unadjusted standard deviation (OR). We then adjusted for age and BMI as linear terms and refitted the unadjusted measures on their own and in combination, as in Table 2 where we estimated OPERAs using the adjusted measures and standard deviations. As before, we compared model fits (log likelihoods and changes in log likelihoods) and conclusions. In particular, we compared the information on risk gradient by fitting a single parameter between that using OPERA with that using OR by comparing Z_OPERA_ = log(OPERA) divided by the standard error of log(OPERA) with Z_OR_ = log(OR) divided by the standard error of log(OR).

**Table 2.**
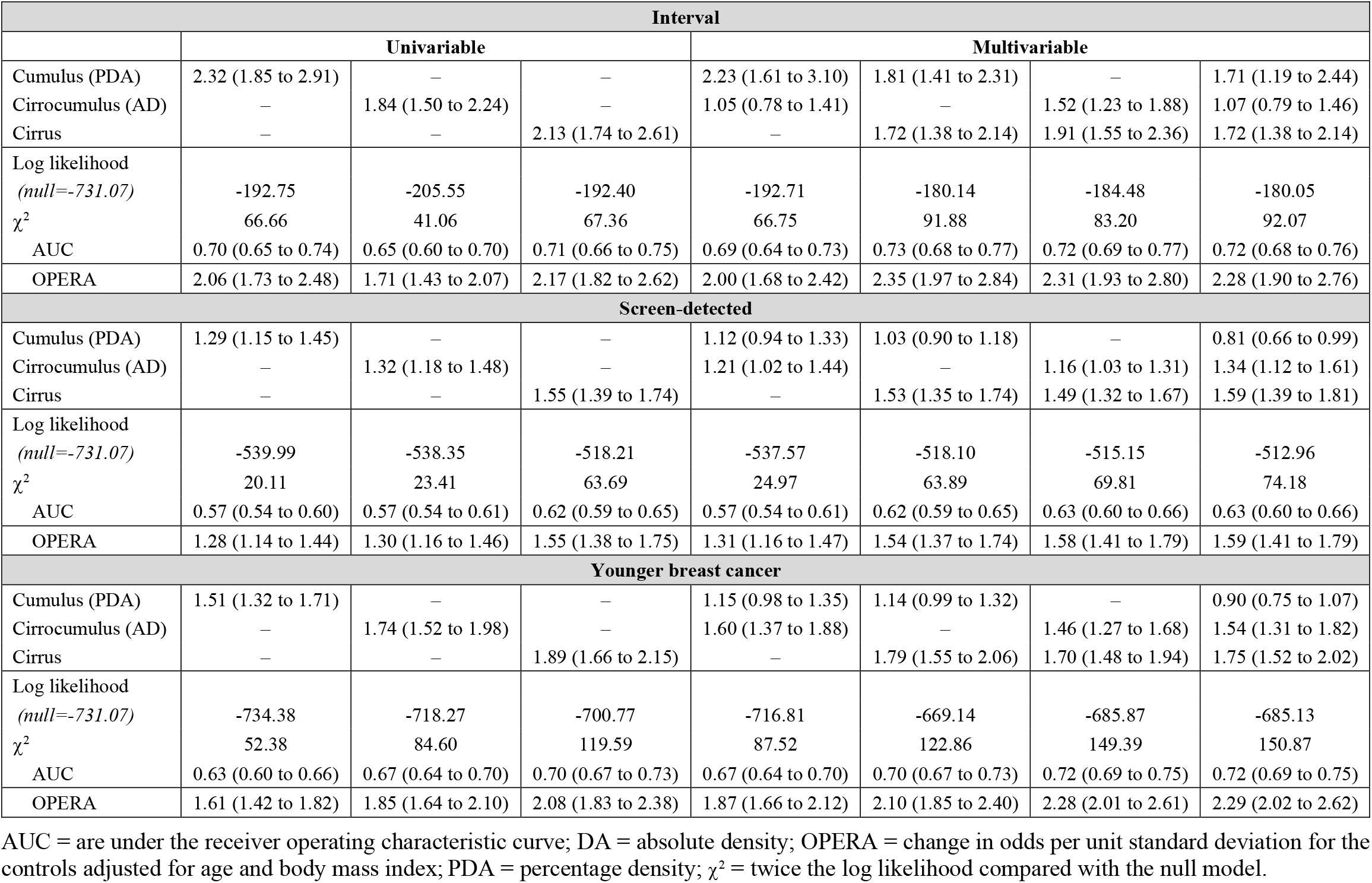
OPERA (95% CI) estimates of odds ratio per adjusted standard deviation from univariable and multivariable analyses of *Cumulus* (as a percentage), *Cirrocumulus* and *Cirrus*, all normalised, adjusted for age and body mass index, and standardised.

## RESULTS

Combining data from the three control samples, the correlation between *Cirrocumulus* and *Cirrus* was 0.3, while the correlations of percentage *Cumulus* with *Cirrocumulus* and *Cirrus* were both 0.5; see Supplementary Figures. The standard errors of these correlations were ∼0.02.

### Interval breast cancer

Table 1 shows that when *Cumulus* and *Cirrus*, or *Cirrocumulus* and *Cirrus*, were fitted together, their risk associations both attenuated but remained significant. When all three were fitted together, *Cirrocumulus* was not significant (*P*=0.6). The best-fitting model involved *Cumulus* and *Cirrus* with log(OPERA)s of 0.59 (95% confidence interval [CI] 0.34 to 0.84) and 0.54 (95% CI, 0.32 to 0.76), respectively. The AUC-equivalent log(OPERA) was 0.85 (95% CI, 0.68 to 1.04). The AUC of 0.73 for the best-fitting model was greater than the AUC of 0.70 for fitting *Cumulus* alone (*P*=10^−6^).

We estimated that women in the highest tertiles of both *Cumulus* and *Cirrus* are at 2.51 (95% CI, 1.72 to 3.64) times the population average risk (*P*<0.001), and this group comprised 17% of the relevant population. At the other extreme, women in the lowest tertiles of both *Cumulus* and *Cirrus* are at 0.22 (95% CI, 0.08 to 0.49) times the population average risk (*P*<0.001), and this group comprised 19% of the population; see Supplementary Table 1. Similar findings were obtained when stratifying women by tertiles of both *Cirrocumulus* and *Cirrus*; see Table 2.

### Screen-detected breast cancer

Table 3 shows that, when *Cumulus* was included with *Cirrus* or *Cirrocumulus*, there was no improvement in fit and the *Cumulus* estimate was no longer significant. When *Cirrocumulus* and *Cirrus* were fitted together, their risk associations both attenuated but remained significant. The best fitting model involved *Cirrus* and *Cirrocumulus* with log(OPERA)s of 0.40 (95% CI, 0.28 to 0.51) and 0.15 (95% CI, 0.03 to 0.27), respectively. For the combined measures, the AUC-equivalent log(OPERA) was 0.46 (95% CI, 0.34 to 0.58). The AUC of 0.63 for the best-fitting model was greater than the AUC of 0.57 for fitting *Cumulus* alone (*P*=10^−12^).

**Table 3.** For *Cirrus* and *Cirrocumulus*, risk relative to the population average, with 95% CI in parentheses, and numbers of cases and controls as a ratio, by tertile-by-tertile.

|  |  | *Cirrus* |  |  |  |  |  |  |  |  |
| --- | --- | --- | --- | --- | --- | --- | --- | --- | --- | --- |
| *Cirrocumulus* |  | Interval |  |  | Screen-detected |  |  | Younger-Diagnosis |  |  |
|  |  | low |  | high | low |  | high | low |  | high |
| low |  | 0.27<br>(0.10 to 0.59) | 0.59<br>(0.26 to 1.22) | 1.17<br>(0.58 to 2.24) | 0.50<br>(0.32 to 0.75) | 0.58<br>(0.37 to 0.91) | 1.42<br>(0.97 to 2.06) | 0.25<br>(0.13 to 0.44) | 0.35<br>(0.17 to 0.66) | 0.53<br>(0.28 to 0.95) |
|  |  | 7/78 | 10/50 | 15/38 | 30/170 | 27/131 | 49/98 | 14/149 | 12/91 | 15/75 |
|  |  | 0.20<br>(0.05 to 0.53) | 0.83<br>(0.43 to 1.52) | 1.45<br>(0.81 to 2.52) | 0.82<br>(0.55 to 1.18) | 0.76<br>(0.50 to 1.14) | 1.22<br>(0.85 to 1.72) | 0.73<br>(0.46 to 1.13) | 0.85<br>(0.56 to 1.27) | 1.89<br>(1.33 to 2.69) |
|  |  | 4/62 | 16/57 | 23/47 | 42/146 | 34/127 | 54/126 | 29/106 | 37/116 | 66/93 |
| high |  | 0.46<br>(0.11 to 1.34) | 1.11<br>(0.62 to 1.90) | 2.45<br>(1.67 to 3.60) | 0.58<br>(0.32 to 1.00) | 0.97<br>(0.67 to 1.38) | 1.96<br>(1.50 to 2.55) | 0.84<br>(0.47 to 1.46) | 0.69<br>(0.43 to 1.08) | 2.45<br>(1.86 to 3.21) |
|  |  | 4/26 | 22/59 | 67/81 | 17/83 | 48/141 | 121/175 | 19/60 | 28/108 | 134/146 |

We estimated that women in the highest tertiles of both *Cirrocumulus* and *Cirrus* are at 2.01 (95% CI, 1.54 to 2.61) times the population average risk (*P*<0.001), and this group comprised 15% of the relevant population. At the other extreme, women in the lowest tertiles of both *Cirrocumulus* and *Cirrus* are at 0.52 (95% CI, 0.33 to 0.78) times the population average risk (*P*<0.001), and this group comprised 14% of the relevant; see Table 2.

### Younger-diagnosis breast cancer

Table 4 shows that, when fitted alone, there was very strong evidence that the model including *Cirrus* had the best fit, and the fit was improved when further including *Cirrocumulus* (*P*<0.001). The addition of *Cumulus* did not improve the fit (*P*=0.8). The best-fitting model involved *Cirrus* and *Cirrocumulus* with log(OPERA)s of 0.53 (95% CI, 0.39 to 0.66) and 0.38 (95% CI, 0.24 to 0.52), respectively. The AUC-equivalent log(OPERA) was 0.82 (95% CI, 0.70 to 0.96). The AUC of 0.72 for the best-fitting model was greater than the AUC of 0.63 for fitting *Cumulus* alone (*P*<10^−23^).

We estimated that women in the highest tertiles of both *Cirrocumulus* and *Cirrus* are at 2.54 (95% CI, 1.93 to 3.33) times the population average risk (*P*<0.001), and this group comprised 16% of the relevant population. At the other extreme, women in the lowest tertiles of both *Cirrocumulus* and *Cirrus* are at 0.39 (95% CI, 0.24 to 0.62) times the population average risk (*P*<0.001), and this group comprised 17% of the population; see Table 2.

Figure 1 shows that the receiver operating curves for *Cirrus* and *Cirrocumulus* have different shapes and crossed over. For *Cirrus*, the sensitivity increased rapidly from zero as the specificity decreased from 1, while for *Cirrocumulus*, the specificity increased rapidly from zero as the sensitivity decreased from 1.

**Figure 1:** Receiver operating characteristic (ROC) curves for *Cirrocumulus* and *Cirrus* for case-control study of younger-diagnosis cancer.

There was no evidence that the OPERAs for *Cumulus, Cirrocumulus* or *Cirrocumulus* differed by age, BMI, menopausal status or family history for any measure within any study (all *P*>0.05). For *Cumulus*, the estimate was greater for interval cancer than both screen-detected and younger-diagnosis breast cancer (*P*=0.00001 and *P*=0.001, respectively). For *Cirrocumulus*, the estimate was greater for younger-diagnosis than for screen-detected breast cancer (*P*=0.002). For *Cirrocumulu*s and *Cirrus*, both estimates were greater for interval than for screen-detected breast cancer (*P*=0.005 and 0.007, respectively).

### Comparison analyses

Supplemental Tables 2 and 3 present analyses for pre- and post-menopausal women, respectively, and indicate that, as found using OPERA, that the general results do not differ by menopausal status.

Combining pre- and post-menopausal women, for interval breast cancer, the OR per unadjusted standard deviation for Percent Density (unadjusted conventional mammographic density as a percentage) was 2.03 (1.63 to 2.53) with Z_OR_ = 6.31. After adjustment for age and BMI, the OR for per unadjusted standard deviation for adjusted Percent Density became 2.67 (95% CI 2.04 to 3.49) (39% greater on the log scale) with Z_OPERA_ = 7.18. For Percent Density, the OPERA was 2.32 (95% CI 1.85 to 2.91) with Z_OPERA_ = 7.27. That is, the OR is much larger than the OPERA even though they are essentially referring to the same concept and, by comparing the Z scores and changes in log likelihood, they are capturing the same amount of information about risk; see Discussion..

Addition of Cirrocumulus and Cirrus gave improved fits with similar changes in twice the log likelihood to what was observed when using OPERA (43.4 cf. 43.8, and 0.2 cf. 0.1, respectively). The best fitting model involved Percent Density and Cirrus.

For screen-detected breast cancer, the OR per unadjusted standard deviation for Percent Density was 1.14 (95% CI 1.02 to 1.28) with Z_OR_ = 2.26. After adjustment for age and BMI it became 1.38 (95% CI 1.21 to 1.58), almost 2.5 times greater on the log scale, with Z_OR_ = 4.73. The OPERA for Percent Density was 1.29 (95% CI 1.15 to 1.45) with Z_OPERA_ = 4.31. Addition of Cirrocumulus and Cirrus gave improved fits with similar changes in twice the log likelihood to what was observed using OPERA (26.6 cf. 25.2, and 5.5 cf. 4.9, respectively). After adjusting for Cirrus and/or Cirrocumulus, the Percent Density estimate was no longer significant, and the best fitting model involved Cirrus and Cirrocumulus.

For younger-diagnosis breast cancer, the OR per unadjusted standard deviation for Breast Density was 1.55 (95% CI 1.36 to 1.76); Z_OR_ = 6.66. After adjustment for age and BMI it became 1.58 (95% CI 1.36 to 1.84), Z_OR_ = 5.93. The OPERA for Cumulus was 1.51 (95% CI 1.32 to 1.71), with Z_OPERA_ = 6.24. Addition of Cirrocumulus and Cirrus gave improved fits with similar changes in twice the log likelihood to what was observed using OPERA (68.56 cf. 70.48, and 35.1 cf. 35.1, respectively). After adjusting for Cirrus and/or Cirrocumulus, the Cumulus estimate was no longer significant, and the best fitting model involved Cirrus and Cirrocumulus.

Therefore, in terms of information about risk, comparisons of Z_OR_ with Z_OPERA_ show that effectively the same models are being fitted once adjustments are made for age and BMI, but the changes in model fits are not always properly captured when the OR estimate is interpreted without taking into account its imprecision.

## DISCUSSION

Our new mammogram-based risk measures based on brightness (*Cirrocumulus*) and texture (*Cirrus*) improved risk prediction beyond an established measure of mammographic density (*Cumulus*). For screen-detected breast cancers and on average younger-diagnosis breast cancers, the best fitting models included both the new measures and performed substantially better than using the established measure alone by doubled the risk gradient (*P*<10^−12^). Except for interval cancers, the new measures also negated the importance of the established measure, *Cumulus*, on risk prediction.

We also found that, when combined, the new mammogram-based risk measures are at least as powerful in identifying women who will be diagnosed with breast cancer as the recently published polygenic risk score [15] which has an OPERA of ∼1.6 [15] or log(OPERA) = 0.48. For younger-diagnosis breast cancer, the AUC for the combination of our measures was 0.72, so the AUC-equivalent log(OPERA) was 0.82. Therefore, in terms of differentiating women with or without breast cancer at a young age, our measures have ([0.82−0.48]/0.48)×100 = 70% more discriminatory power than does the polygenic risk score. It is plausible that inclusion of a polygenic risk score with the mammogram-based risk measures will further improve risk prediction [16].

On a population basis, therefore, the combination of these new measures appears to be the strongest of all known breast cancer risk factors. For example, when *Cirrocumulus* and *Cirrus* were combined to predict breast cancer at on average a younger age (see Table 1), the AUC-equivalent OPERA was 2.28, so the interquartile risk ratio is ∼7-fold. In comparison, the interquartile risk ratio is ∼4-fold for a multigenerational family history risk score in predicting breast cancer before age 50 years, ∼3-fold for the latest polygenic risk score, ∼2-fold for conventional mammographic density, ∼1.5-fold for *BRCA1* and *BRCA2* mutations, and ∼1.2-fold or less for lifestyle-related risk factors [7,8]..

From the contrasting shapes of their receiver operating characteristic curves, it can be seen that *Cirrus* has greater sensitivity at high specificity (i.e. when better identifying true negatives), while *Cirrocumulus* has greater specificity at high sensitivity (i.e. when better identifying true positives). Therefore, *Cirrocumulus* does better at identifying women at higher than average risk, while *Cirrus* does better at identifying women at lower than average risk.

*Cirrus* gave similar risk gradients in all three settings, suggesting it is tapping into new and fundamental risk-predicting aspects of a mammogram. This was also evident in the original work developing *Cirrus*, which found that a similar risk prediction was achieved for women of Japanese ancestry living in Hawaii and for Australian women [6]. Note that *Cirrus* was designed not to depend on brightness and has only a modest correlation with the brightness measures.

The same general conclusions about best fitting models were observed when we used the OPERA approach as when we used the standard approach; see Comparison analyses in the Results. However, the latter is problematic because the OR per unadjusted standard deviation is prone to be misinterpreted when it is estimated from fitting a model that includes factors associated with the measure. This was clearly evident when fitting Percent Density and Cirrus as predictors of interval breast cancer; see Results.

Our new mammogram-based measures are potentially of substantial clinical and population health significance. They not only identify groups of women at substantially increased risk, but they also identify larger groups of women at decreased risk. When categorised by tertiles, *Cirrus* and *Cirrocumulus* divide the population into two extreme groups of approximately the same size (each about 15–20%) containing women who are on average either at twice or more population risk, or at half or less population risk; see Table 2. For interval cancer, about 60% of controls were in the six categories with below population average risk, while for both screen-detected and interval cancer, about 75% of controls were in the six categories with below population average risk.

These observations are highly relevant to considerations of tailored, or personalised, screening based on risk, for which there are now several trials being conducted across the world. These include the Wisdom Study in the United States [17,18], My personalised breast screening (MyPeBS) in France (https://clinicaltrials.gov/ct2/show/NCT03672331), and PROCAS2 in the United Kingdom (https://preventbreastcancer.org.uk/breast-cancer-research/research-projects/early-detection-screening/procas/).

These risk categorisations are in stark contrast to those using BI-RADS alone. Currently, about 40% of screening women in the United States are classified as having dense breasts defined by BI-RADS categories c or d. As a result of a community-led initiative [19], in 35 states it is mandated by law that these women are notified. Research studies in which one or a few radiologists measure BI-RADS in a controlled manner suggest the increased risk associated with having dense breasts is about 1.6 to 2.2-fold (see IBIS [20] and BOADICEA [21]).

In practice, BI-RADS is measured by multiple radiologists at a given screening service, especially over time, opening the potential for substantial measurement error. For example, from the Supplemental data on 60,000 women screened at a large United States medical centre [22], the odds ratio for breast cancer based on being classified as having dense breasts is only about 1.1, which is far less than the typical odds ratios found by research studies (*P*<0.001). This was despite the measurements being recorded by “radiologists who specialized in breast imaging and who had 5–33 years of experience following the American College of Radiology BI-RADS lexicon” [23]. It would appear, therefore, that in practice there could be so much variation across measurers, even experienced specialists in a large city-based service, that clinical BI-RADS measurements might be providing very little information on risk stratification across the population.

There is substantial scope for better addressing the issue of dense breasts by going beyond BI-RADS. A major consequence of having dense breasts is an increased risk of interval cancer. We and others have found that, as well as conventional mammographic density (*Cumulus*), having a family history and other risk factors, such as cumulative exposure to ovarian hormones based on the Pike model [24], combine to predict interval cancer [11]. In our study we have found that *Cirrus* also brings almost as much information as *Cumulus*, and when combined they have an inter-quartile risk ratio for interval cancer of almost 9-fold. Future work will consider how risk of interval cancer, and even of missed cancers, can be further optimised by combining mammogram-based measures with family history, genetic risk scores and other risk factors. This could have a profound impact on the way the issue of dense breasts is addressed in the future.

For our findings to be translated into wider clinical practice, automated use of the mammogram-based and other risk measures needs to be implemented. We are developing a program to measure *Cirrus* automatically from batch files of digital or digitised mammograms and are using deep learning to develop similar automated measures of *Cumulus* and *Cirrocumulus*. We are developing the empirical evidence, such as in this and other papers [11, 16] to find out how mammogram-based risk measures combine with each other and with other important risk factors to predict risk.

In conclusion, the established mammographic density measure improved the prediction of interval cancers, most likely due to its role in masking tumours. But this measure provided no substantial, additional risk information on top of our new mammogram-based risk measures for screen-detected or younger-diagnosis cancer. Therefore, conventional mammographic density appears to cause existing tumours to be missed, but not necessarily to cause breast tumours to develop in the first place. There are likely to be other aspects of woman’s breasts that are detectable from a mammogram and have a truly causal effect on breast cancer initiation and progression. Our new measures appear to be more strongly correlated with such causal factors than conventional mammographic density. Our findings also demonstrate the potential for much improved and more aetiologically relevant breast cancer risk prediction, by discovering new ways of extracting information on breast cancer risk from a mammogram. This suggests a way that risk-based personalised breast screening could become part of the precision medicine era [17, 18].

### Consent for publication

Not applicable.

## Data Availability

For data accessment, please contact the corresponding author.

## Acknowledgements

We thank the Breast Cancer Network Australia, the Victorian Cancer Registry, BreastScreen Victoria, the Australian Mammographic Density Research Facility, and the participants in the Melbourne Collaborative Cohort Study.

## Dedication

This paper is dedicated to the memory of Nancy Cappello and her achievements in raising awareness of the role of breast density in masking breast tumors.

## AUTHOR CONTRIBUTIONS

*Study design:* TLN, DFS, EM, GM, SL, GSD, YMS, JSu, LB, MAJ, RLM, GGG, JLH. *Data collection and preparation:* TLN, DFS, EM, GM, SL, YKA, CFE, HNT, LB, JSt, PAD, MCS, GGG, MAJ, RLM, JLH. *Data analysis:* TLN, DFS, EM, GM, JLH. *Data interpretation:* TLN, DFS, EM, GM, SL, GSD, YKA, CFE, HNT, LB, JSt, YMS, JSu, RJM, PAD, JGD, MAJ, RLM, MCS, GGG, JLH. *Manuscript drafting:* TLN, DFS, EM, JLH. All author contributed to the preparation and approved the final version of manuscript.

## Ethics Statement

All participants gave written informed consent and the studies were approved by appropriate human research ethics committees.

## Data Accessibility

The datasets used for the current study are available upon reasonable request from the corresponding author and with permission of appropriate human research ethics committees.

A preprint version of this manuscript has been posted on the medRxiv with the DOI: https://doi.org/10.1101/2020.05.24.20111815 (https://www.medrxiv.org/content/10.1101/2020.05.24.20111815v1)

## Abbreviations

AUC: Area under the receiver operating characteristic curve;
BMI: Body mass index;
CI: Confidence interval;
CC: Cranio-caudal;
LL: log likelihood;
OPERA: odds per adjusted standard deviation.

## REFERENCES

1. Boyd NF, Guo H, Martin LJ, et al. Mammographic density and the risk and detection of breast cancer. N Engl J Med 2007;356:227–36.

2. Nguyen TL, Aung YK, Evans CF, et al. Mammographic density defined by higher than conventional brightness threshold better predicts breast cancer risk for full-field digital mammograms. Breast Cancer Res 2015;17:142.

3. Nguyen TL, Aung YK, Evans CF, et al. Mammographic density defined by higher than conventional brightness thresholds better predicts breast cancer risk. Int J Epidemiol 2017;46:652–661.

4. Nguyen TL, Choi YH, Aung YK, et al. Breast cancer risk associations with digital mammographic density by pixel brightness threshold and mammographic system. Radiology 2018;286:433–442.

5. Nguyen TL, Aung YK, Li S, et al. Predicting interval and screen-detected breast cancers from mammographic density defined by different brightness thresholds. Breast Cancer Res 2018;20:152.

6. Schmidt DF, Makalic E, Goudey B, et al. Cirrus: an automated mammography-based measure of breast cancer risk based on textural features. JNCI Cancer Spectrum 2018;2(4).pky057.

7. Hopper JL, Nguyen TL, Schmidt DF, et al. Going beyond conventional mammographic density to discover novel mammogram-based predictors of breast cancer risk. J Clin Med 2020; 9:(3).pii: E627.

8. Hopper JL. Odds per adjusted standard deviation: comparing strengths of associations for risk factors measured on different scales and across diseases and populations. Am J Epidemiol 2015;182:863–867.

9. Baglietto L, Krishnan K, Stone J, et al. Associations of mammographic dense and nondense areas and body mass index with risk of breast cancer. Am J Epidemiol 2014;179:475–83.

10. Krishnan K, Baglietto L, Apicella C, et al. Mammographic density and risk of breast cancer by mode of detection and tumor size: a case-control study. Breast Cancer Res 2016;18:63.

11. Nguyen TL, Li S, Dite GS, et al. Interval breast cancer risk associations with breast density, family history, and breast tissue ageing. Int J Cancer 2019;145:1768–1773.

12. Byng JW, Yaffe MJ, Jong RA, et al. Analysis of mammographic density and breast cancer risk from digitized mammograms. Radiographics 1998;18:1587–98.

13. StataCorp. Stata statistical software: Release 15 College Station, TX: StataCorp LLC, 2017.

14. Fisher RA. Statistical methods and scientific induction. J Roy Statist Soc B 1995;17:69–78.

15. Mavaddat N, Michailidou K, Dennis J, et al. Polygenic risk scores for prediction of breast cancer and breast cancer subtypes. Am J Hum Genet 2019;104:21–34.

16. Vachon CM, Scott CG, Tamimi RM, et al. Joint association of mammographic density adjusted for age and body mass index and polygenic risk score with breast cancer risk. Breast Cancer Res 2019;21:68.

17. Esserman LJ, and the WISDOM Study and Athena Investigators. The WISDOM Study: breaking the deadlock in the breast cancer screening debate. NPJ Breast Cancer 2017;3;34.

18. Shieh Y, Eklund M, Madlensky L, et al. Breast cancer screening in the precision medicine era: risk-based screening in a population-based trial. J Natl Cancer Inst. 2017;109(5): doi:10.1093/jnci/djw290.

19. Cappello NM, Richetelli D, Lee CI. The impact of breast density reporting laws on women’s awareness of density-associated risks and conversations regarding supplemental screening with providers. J Am Coll Radiol 2019;16:139–46.

20. Warwick J, Birke H, Stone J, et al. Mammographic breast density refines Tyrer-Cuzick estimates of breast cancer risk in high-risk women: findings from the placebo arm of the International Breast Cancer Intervention Study I. Breast Cancer Research 2014;16:451–6.

21. Lee A, Mavaddat N, Wilcox AN, et al. BOADICEA: a comprehensive breast cancer risk prediction model incorporating genetic and nongenetic risk factors. Genetics in Medicine 2019;21:1708–1718.

22. Yala A, Lehman C, Schuster T, et al. A Deep Learning Mammography-based Model for Improved Breast Cancer Risk Prediction. Radiology 2019;291:60–66.

23. Lehman CD, Yala A, Schuster T, et al. Mammographic breast density assessment using deep learning: clincal implementation. Radiology 2019;290:52–58.

24. Pike MC, Krailo MD, Henderson BE, Casagrande JT, Hoel DG. ‘Hormonal’ risk factors, ‘breast tissue age’ and the age-incidence of breast cancer. Nature 1983;303:767–70.

